# CRISPR-Cas3-Based Detection of *Cryptosporidium* and Pepper Mild Mottle Virus in Environmental Waters Using a Test-Strip Readout

**DOI:** 10.64898/2026.11.13.26368066

**Authors:** Shotaro Torii, Rika Hirano, Made Sandhyana Angga, Kohei Takeshita, Kazuto Yoshimi, Masaaki Kitajima, Tomoji Mashimo, Kumiko Oguma

## Abstract

Test-strip-based CRISPR diagnostics could enable decentralized nucleic acid monitoring, but their application to environmental waters has been constrained by insufficient sensitivity and complex environmental matrices. Here, we developed a highly sensitive and specific assay combining reverse transcription-nested recombinase polymerase amplification (RT-nRPA) with Cas3-Operated Nucleic Acid detectioN (CONAN) for test-strip-based detection of *Cryptosporidium* 18S rRNA and pepper mild mottle virus (PMMoV). Introducing a second, nRPA step reduced the 50% limit of detection (LOD_50_) to 2.0 copies/reaction for *Cryptosporidium* and 1.5 copies/reaction for PMMoV. In 17 river water and six wastewater extracts, CONAN was positive for all nRPA products containing the *Cryptosporidium* sequence and negative for all products lacking the target. Cas12a-based DETECTR was negative for four *Cryptosporidium*-containing nRPA products, whereas LAMP and qPCR frequently cross-reacted with non-target environmental eukaryotic sequences. Consequently, nRPA-CONAN can overcome sensitivity and specificity constraints and offer a practical route toward instrument-light nucleic acid monitoring in environmental waters.

## 1. Introduction

Nucleic acid biomarkers are increasingly used for pathogen monitoring in drinking water sources and as a tool for wastewater-based epidemiology (WBE). Despite the growing demand for such applications, nucleic acid detection still primarily relies on quantitative PCR (qPCR), which requires a thermal cycler equipped with a fluorescence reader as well as trained personnel^1^. Isothermal amplification methods, such as loop-mediated isothermal amplification (LAMP)^2^ and recombinase polymerase amplification (RPA)^3^, offer a simpler temperature control, but they can generate nonspecific amplification products from closely related sequences or primer-derived products.^4–6^ Thus, there is a need for sensitive, sequence-specific molecular assays that can operate with minimal instrumentation^7^.

Clustered regularly interspaced short palindromic repeats (CRISPR) diagnostics have the potential to address these unmet needs. In CRISPR systems, CRISPR RNA (crRNA) guides CRISPR-associated (Cas) proteins to specific nucleic acid targets, and these systems have been repurposed for diagnostic applications^8^ and increasingly applied to water testing^4,9–11^. Among the available signal-readout formats for CRISPR diagnostics, lateral flow assay (LFA) is one of the most common and instrument-light formats because it provides a visible test-strip-based readout. One widely used example is DNA endonuclease-targeted CRISPR trans reporter (DETECTR), which couples isothermal amplification with an additional sequence-recognition step by Cas12a-crRNA complex recognizing a 20-base sequence downstream of protospacer adjacent motif (PAM)^12^. Another emerging example is Cas3-Operated Nucleic Acid detectioN (CONAN), targeting a longer 32-base sequence with a stringent PAM recognition^13^. This extended recognition may increase discrimination among closely related sequences. Both DETECTR and CONAN operate at approximately 37 °C and are therefore compatible with RPA. However, to our knowledge, these two CRISPR diagnostic platforms have not been directly benchmarked against conventional molecular assays, such as qPCR and LAMP, in environmental water samples.

In contrast to specificity, for CRISPR diagnostics, achieving analytical detection limits in the few-copies-per-reaction range has not always been a primary focus, particularly for clinical applications, in which positive samples may contain target concentrations well above the detection threshold.^14^ In such settings, modest differences in analytical sensitivity may have limited effects on qualitative classification, and greater emphasis has thus been placed on simplicity, rapidity, and portability. For example, reported LODs for RPA-DETECTR range from tens to thousands of copies per µL with LFA.^15^ Similarly, previous LFA-based RPA-CONAN assays have required hundreds of target copies per reaction^16^. In environmental waters, however, target concentrations can be substantially lower, making high analytical sensitivity a critical requirement for extending CRISPR diagnostics to water monitoring.

Here, we developed a sensitive and specific LFA-based CONAN platform for detecting *Cryptosporidium* 18S rRNA and pepper mild mottle virus (PMMoV) in environmental waters. *Cryptosporidium* is an important waterborne protozoan parasite transmitted through contaminated drinking and recreational waters^17^, whereas PMMoV is widely used as an indicator of fecal contamination and an endogenous process control^18^. We first identified insufficient amplification by conventional RPA as a major factor limiting the analytical sensitivity of LFA-based detection and therefore developed a nested RPA (nRPA) strategy to achieve detection at a few copies per reaction. We then applied the optimized assay to 17 river-water and six wastewater extracts and benchmarked the performance of nRPA-CONAN against nRPA-DETECTR, LAMP, and qPCR, using nanopore sequencing of the corresponding amplicons as reference for verifying the target.

## 2. Results

### 2.1. Working principle of CONAN and assay design for the detection of *Cryptosporidium* 18S rRNA and PMMoV

The working principle of CONAN is shown in Figure 1. In CONAN, the target is recognized by the Cascade (CRISPR-associated complex for antiviral defense)–crRNA complex. Target recognition requires a PAM, such as 5’-AAG-3’^13^. Any sequences adjacent to the PAM can be targeted by reprogramming the crRNA sequence. Upon sequence-specific binding of the Cascade–crRNA complex to the target DNA, Cas3 is recruited and activated, triggering non-specific cleavage of single-stranded DNA (ssDNA) (i.e., collateral activity). This activity can be quantified using exogenously added fluorophore–quencher ssDNA probes (FQ probes), which generate increased fluorescence upon cleavage. For LFA, fluorescein–biotin ssDNA probes (FB probes) can instead be used to provide a visual readout.^19^ (Figure 1).

**Figure 1.**
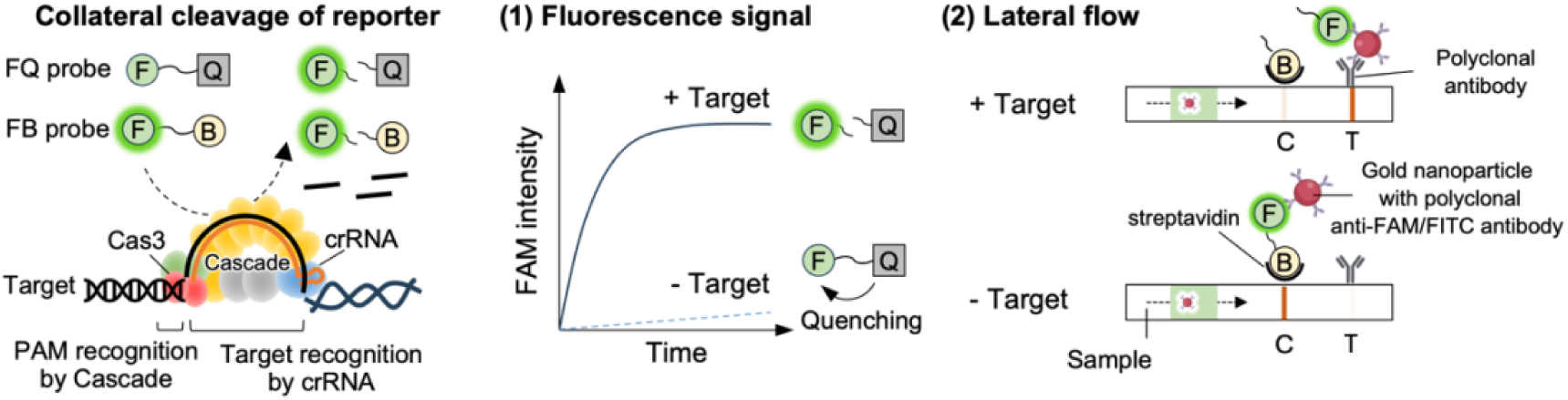
Working principle of CONAN. Upon sequence-specific binding of the Cascade–crRNA complex to the target DNA, Cas3 is recruited and activated, triggering collateral cleavage of single-stranded DNA reporters. Cleavage of fluorophore– quencher (FQ) reporters produces a fluorescence signal (1), whereas cleavage of FAM– biotin (FB) reporters is visualized by lateral-flow assay (LFA) (2).

For *Cryptosporidium*, 18S rRNA was selected because of its high intracellular abundance. The rationale is detailed in the Supporting Information (SI). We used primers and crRNAs targeting a conserved region spanning the V3 stem-loop (Table 1 and Table S1, Figure S1), which has frequently been targeted by previous qPCR, LAMP, and RPA assays.^20–25^ The assay was designed primarily for *C. parvum* and *C. hominis*. Two CONAN crRNAs were used as an equimolar mixture to accommodate sequence variation. BLAST-based *in silico* screening showed that the reverse inner primer C18S-R2 and both CONAN crRNAs did not exhibit notable homology to non-*Cryptosporidium* sequences, suggesting that the assay design met a preliminary requirement for *Cryptosporidium*-specific detection. It should be noted that BLAST also showed that the primers and crRNAs are compatible not only with *C. parvum* and *C. hominis* but also with *C. meleagridis* and *C. suis*. The species coverage of the assay was thus evaluated using synthetic DNA (Table S2) as described in the SI. Similarly, for each of the three existing qPCR assays and the LAMP assay evaluated in this study, at least one primer or probe showed no notable homology to non-*Cryptosporidium* sequences, meeting the preliminary requirement.

**Table 1.**
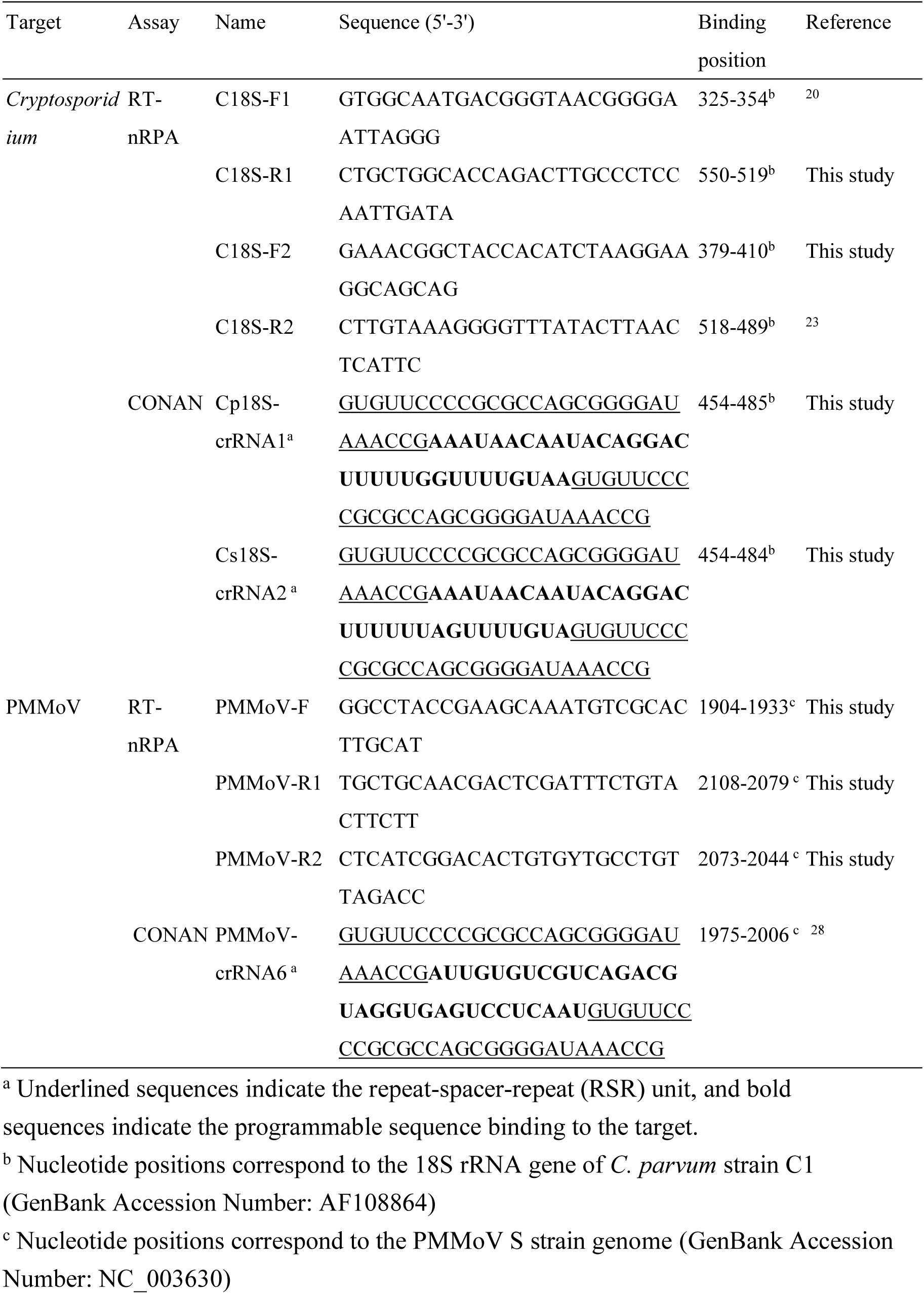
Primers and crRNAs used for RT-nRPA-CONAN detection of *Cryptosporidium*and PMMoV.

For PMMoV, we targeted a region previously used for qPCR^26,27^ and by an RT-LAMP-CONAN assay.^28^ Because sequence variation prevented design of two broadly conserved primer pairs, we used a semi-nested format with a shared forward primer and distinct outer and inner reverse primers. BLAST screening did not identify notable homology of the selected primers or crRNAs to non-PMMoV sequences.

### 2.2. Factor underlying the limited detection of low-copy targets by LFA-based CRISPR diagnostics

To determine why LFA-based CRISPR detection had a relatively high analytical limit of detection, we first compared LFA and fluorescence responses over 5 × 10^8^–5 × 10^11^ copies of *C. parvum* target DNA. Both CONAN and DETECTR produced clear LFA bands at approximately 5 × 10^10^ copies after 10 min (Figure S2A), whereas ≤5 × 10^9^ copies produced no detectable LFA signal despite measurable fluorescence (Figure S2B, C), consistent with a meta-analysis showing a trend of higher analytical LODs of LFA than fluorescence-based detection.^29^ Because 1 µL of amplification product was added to each CRISPR reaction, we used 5 × 10^10^ copies/µL as an empirical concentration threshold for LFA detection.

We next investigated whether conventional RPA could efficiently amplify low initial RNA inputs to the required concentration. Different amounts of *C. parvum* 18S rRNA, ranging from 13 to 1,300 copies per reaction, were reverse transcribed for 10 min in the rehydrated RPA reaction mixture, and then RPA was initiated by the addition of magnesium acetate. Changes in target concentration during 40 min of RPA were quantified (Figure 2) by intercalator dye-based qPCR as detailed in the SI. Rapid amplification occurred during the first 10 min, resulting in an increase of >7 orders of magnitude for all initial RNA inputs. Beyond 10 min, further amplification was limited, with target concentrations increasing by <2 log_10_ thereafter. The 1,300-copy reaction crossed the LFA threshold within 10 min, whereas 13- and 130-copy reactions remained below it after 40 min. Thus, conventional RPA plateaued before low-copy inputs reached the target concentration required for reliable LFA detection, preventing lowering the analytical LOD.

**Figure 2.**
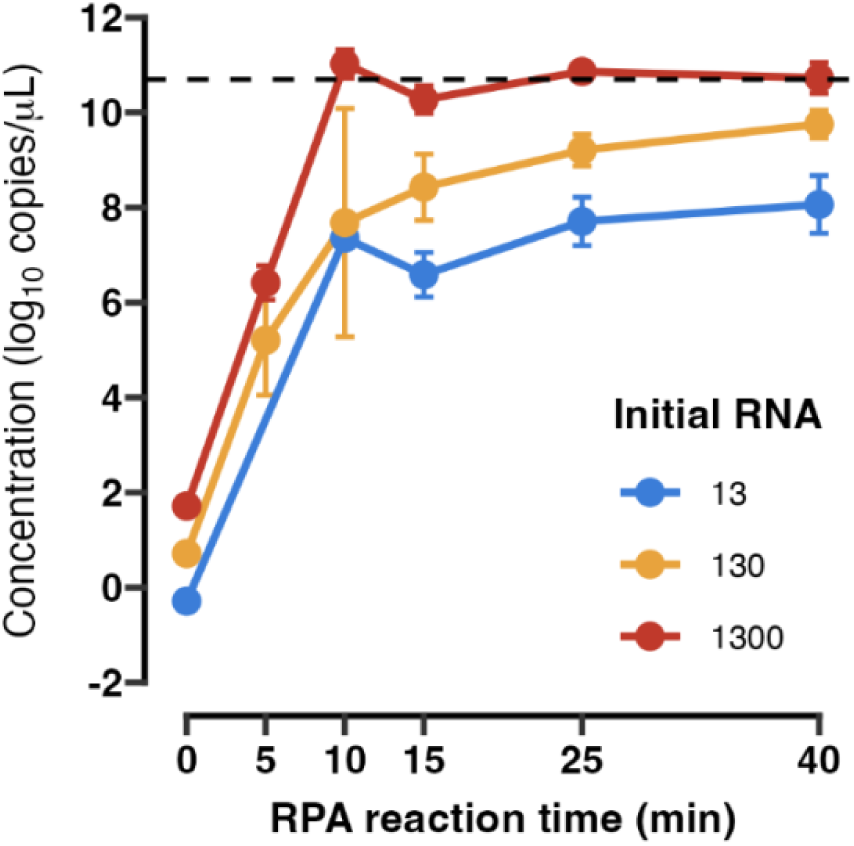
Effect of initial RNA input on RPA amplification kinetics. *C. parvum* 18S rRNA at different initial inputs was reverse-transcribed for 10 min, followed by RPA for 40 min. Target concentrations in the 25-µL reaction mixture were quantified by intercalator dye-based qPCR. Data are shown as mean ± SD. The dashed line represents the empirical concentration threshold for LFA detection, 5×10^10^ copies/μL. Data for the 13-copy input at 5 min are not shown because all measurements were below the lower limit of the qPCR standard curve. Values at 0 min represent the initial RNA copy number divided by the reaction volume (25 µL), corresponding to the estimated concentration at the initiation of RPA following the 10-min reverse-transcription step.

### 2.3. Nested RPA lowers the analytical limit of detection of CONAN

To overcome the amplification plateau of conventional RPA and lower the analytical LOD of the LFA, we developed a nested RPA (nRPA). This approach incorporates a second RPA step in which first-round RPA amplicons are re-amplified using inner primers, thereby enriching the target available for CRISPR detection (Figure 3A). Briefly, RT and nested RPA were performed consecutively in a single tube. RNA was first reverse transcribed for 10 min in the presence of the outer primers, after which magnesium acetate was added to initiate the first RPA reaction for 10 min. A four-fold volume of the second RPA mixture containing the inner primers was then added, and the second RPA reaction was performed for 15 min. The outer primers were used at half the concentration of the inner primers so that, after addition of the second-round mixture, the inner-primer concentration was eightfold higher than that of the outer primers, favoring amplification by the inner primers during the second RPA step.

**Figure 3.**
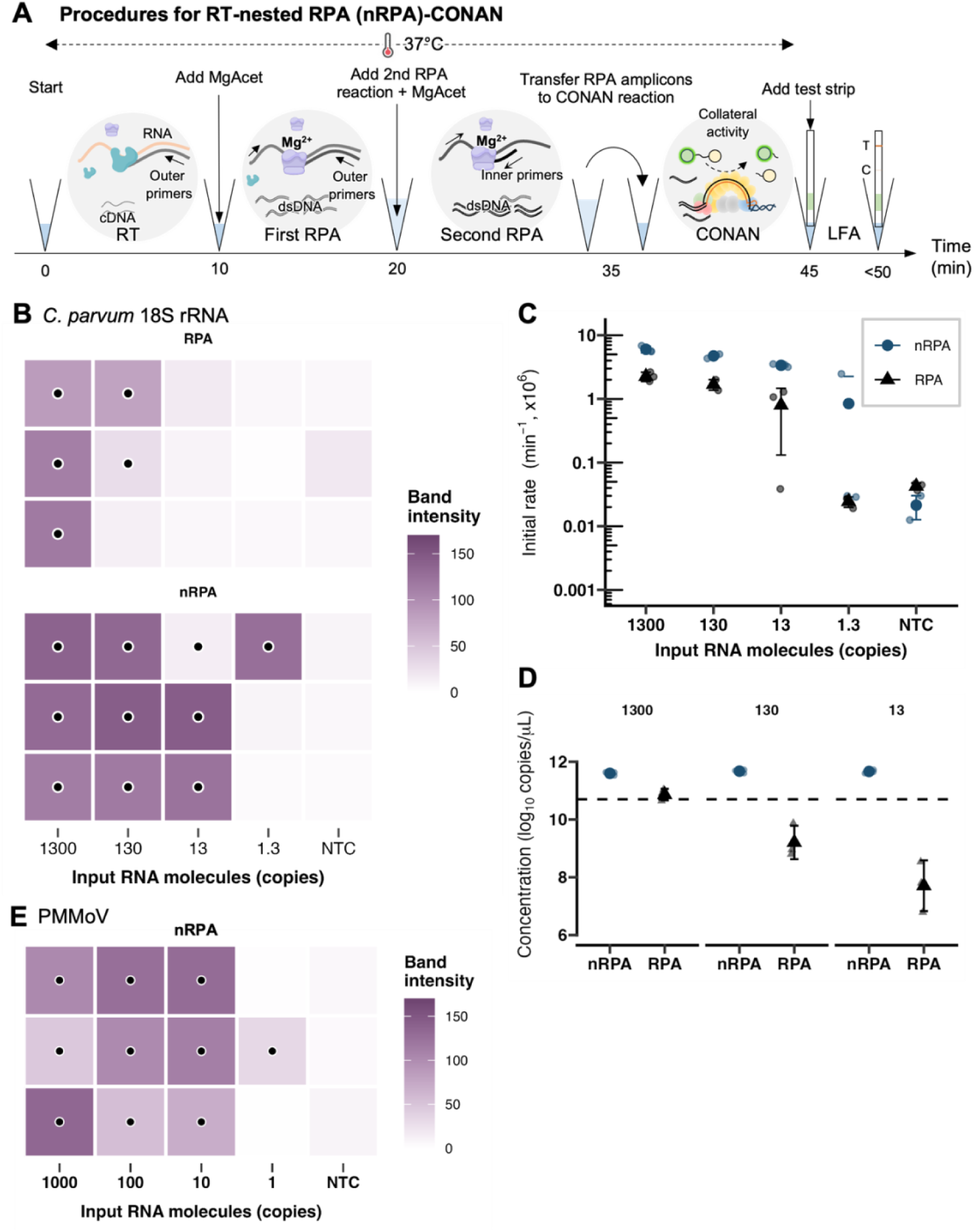
Reduction in the analytical limit of detection (LOD) of CONAN using RT-nRPA. (A) Schematic of the RT-nRPA-CONAN workflow. (B) Heatmaps showing lateral-flow band intensities for different amounts of input *C. parvum* 18S rRNA molecules, 1.3-1,300 copies, with two different procedures, RPA and nRPA. Black dots indicate samples classified as positive, with band intensities exceeding the upper 99% threshold of the non-template controls, calculated as mean + 2.33 standard deviation (SD). (C) Initial fluorescence increase rates. Small symbols indicate individual replicates, and large symbols indicate the mean ± SD. The time-course of fluorescence changes is shown in Figure S3. (D) *C. parvum* target concentrations after nRPA at different initial RNA inputs, compared with those after 25 min of conventional RPA. Concentrations in the 25 μL reaction mixture were quantified by intercalating dye-based qPCR. Data are shown as mean ± SD. The data for RPA were taken from Figure 2 for reference. The dashed line represents the estimated concentration required for LFA detection, 5×10^10^ copies/μL. (E) Heatmaps showing lateral-flow band intensities for different amounts of input PMMoV RNA molecules, 1-1,000 copies, for RT-nRPA-CONAN. Black dots indicate samples classified as positive, with band intensities exceeding the upper 99% threshold of the non-template controls, calculated as mean + 2.33 SD.

To experimentally evaluate the nRPA approach, we compared the analytical LOD of CONAN following conventional RPA and nRPA. LFA test band intensities, initial fluorescence increase rates of the CONAN reaction, and the target concentrations after amplification are shown in Figure 3B-D, respectively. For all tested initial RNA inputs, nRPA yielded higher target concentrations than conventional RPA, reaching 11.6 to 11.7 log_10_ copies/μL and exceeding the concentration required for LFA-based detection (Figure 3D). Consistent with these results, nRPA produced stronger LFA test-bands (Figure 3B) and higher initial fluorescence increase rates in the CONAN assay (Figure 3C). Notably, the input copy number corresponding to a 50% probability of detection (LOD_50_) in the LFA-based assay decreased from 98 copies/reaction with conventional RPA to 2.0 copies/reaction with nRPA, corresponding to an approximately 49-fold reduction in LOD.

To assess the applicability of the nRPA-CONAN workflow to another RNA target, we applied the nRPA-CONAN workflow to PMMoV. Positive LFA bands were observed even at 1–10 copies/reaction (Figure 3E), with an LOD_50_ of 1.5 copies/reaction. These results demonstrate that nRPA lowers the analytical LOD of CONAN for *Cryptosporidium* and PMMoV to the few-copy-per-reaction range.

### 2.4. Comparison of CONAN specificity with DETECTR, LAMP, and qPCR

Environmental performance was evaluated using 17 river water and six wastewater extracts (Figure 4A and Figure S5). Each extract was analyzed by RT-nRPA-CONAN, RT-nRPA-DETECTR, RT-LAMP^21^, and the three RT-qPCR assays, including qPCR-NJ^22^, qPCR-CM^24^, and qPCR-DW^25^. qPCR-DW yielded no environmental positives despite amplification from positive controls and was thus excluded from comparative analysis. Amplicons from the remaining assays were subjected to nanopore sequencing. The assay and sequencing results are summarized in Table 2 and Table S3, with LFA results presented in Figure 4B and detailed nanopore sequencing results provided in Table S4-S7.

**Figure 4.**
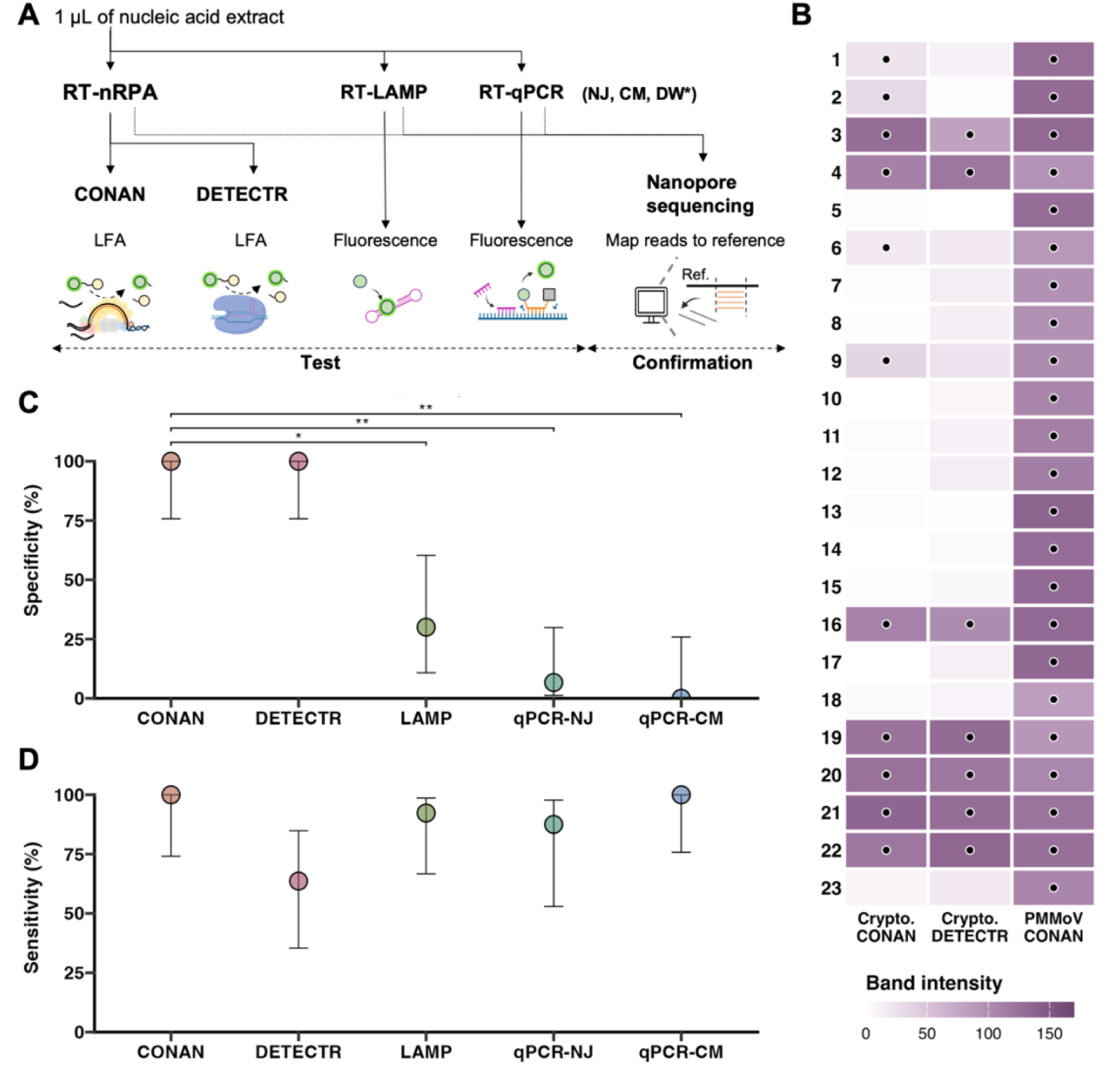
Side-by-side comparison of specificity and sensitivity of CONAN, DETECTR, LAMP, and qPCR for detecting *Cryptosporidium*. (A) Experimental workflow. The asterisk denotes qPCR-DW, which yielded no positive environmental-sample results despite successful amplification of the positive controls and was excluded from subsequent comparative analysis. (B) Heatmap of LFA test-band intensities obtained after nRPA-CONAN, -DETECTR for *Cryptosporidium* (abbreviated as Crypto.) and nRPA-CONAN for PMMoV. Black dots indicate samples classified as positive, with band intensities exceeding the upper 99% threshold of the non-template controls, calculated as the mean + 2.33 SD. (C) Specificity of each assay. Specificity was calculated as TN/(TN + FP), using the presence or absence of *Cryptosporidium*-specific contigs in the corresponding amplification products as the reference. (D) Sensitivity, calculated as TP/(TP + FN) using the same assay-specific reference. Error bars indicate 95% confidence intervals. Brackets indicate the reported pairwise comparisons (McNemar’s test; \**p* < 0.05, \*\**p* < 0.01).

**Table 2.** Detection results for *Cryptosporidium* in environmental water samples. Assay results were compared with the presence or absence of *Cryptosporidium*-specific contigs in the corresponding amplification products. Samples with a positive assay result and *Cryptosporidium*-specific contigs were classified as true positive (TP); samples with a positive assay result but no *Cryptosporidium*-specific contigs were classified as false positive (FP); samples with a negative assay result and no *Cryptosporidium*-specific contigs were classified as true negative (TN); and samples with a negative assay result despite the presence of *Cryptosporidium*-specific contigs were classified as false negative (FN).

| No. | Date | Name | Type | CONAN | DETECTR | LAMP | qPCR-NJ | qPCR-CM |
|---|---|---|---|---|---|---|---|---|
| 1 | 2026/03/23 | Shibuta | River Water | **TP** | FN | **TP** | **TP** | **TP** |
| 2 | 2026/03/23 | Sagami | River Water | **TP** | FN | **TP** | FP | **TP** |
| 3 | 2026/03/23 | Nakatsu | River Water | **TP** | **TP** | **TP** | FP | **TP** |
| 4 | 2026/03/23 | Koayu | River Water | **TP** | **TP** | **TP** | FP | **TP** |
| 5 | 2026/03/23 | Hatsuse | River Water | TN | TN | FP | FP | FP |
| 6 | 2026/04/03 | Shingashi | River Water | **TP** | FN | **TP** | **TP** | **TP** |
| 7 | 2026/04/03 | Shirako | River Water | TN | TN | FP | FP | FP |
| 8 | 2026/04/03 | Kurome | River Water | TN | TN | FP | FP | FP |
| 9 | 2026/04/03 | Arakawa | River Water | **TP** | FN | **TP** | FP | **TP** |
| 10 | 2026/04/03 | Kounuma | River Water | TN | TN | FP | FP | FP |
| 11 | 2026/04/03 | Kamogawa | River Water | TN | TN | FP | FP | FP |
| 12 | 2023/06/14 | Tamagawa | River Water | TN | TN | **TP** | FP | FP |
| 13 | 2023/08/14 | Tamagawa | River Water | TN | TN | FP | FP | FP |
| 14 | 2023/10/03 | Tamagawa | River Water | TN | TN | **TP** | FP | FP |
| 15 | 2023/12/27 | Tamagawa | River Water | TN | TN | TN | FP | FP |
| 16 | 2024/02/08 | Tamagawa | River Water | **TP** | **TP** | **TP** | **TP** | **TP** |
| 17 | 2024/04/12 | Tamagawa | River Water | TN | TN | TN | FP | FP |
| 18 | 2026/02/12 | WWTP-A | Wastewater | TN | TN | FP | TN | FP |
| 19 | 2026/02/19 | WWTP-A | Wastewater | **TP** | **TP** | **TP** | FN | **TP** |
| 20 | 2026/02/10 | WWTP-B | Wastewater | **TP** | **TP** | **TP** | **TP** | **TP** |
| 21 | 2025/04/04 | Airport | Wastewater | **TP** | **TP** | **TP** | **TP** | **TP** |
| 22 | 2025/07/01 | Airport | Wastewater | **TP** | **TP** | FN | **TP** | **TP** |
| 23 | 2025/08/05 | Airport | Wastewater | TN | TN | TN | **TP** | **TP** |

PMMoV was detected by RT-nRPA-CONAN in all 23 samples (Figure 4B), confirming the presence of amplifiable RNA in each extract. PMMoV was also detected by qPCR in all samples, at concentrations of 1.6×10^1^ – 4.3×10^4^ gc/µL in nucleic acid extracts, corresponding to 3.5×10^3^ – 6.5×10^7^ gc/L in water samples. Nanopore sequencing of six selected nRPA products and six selected qPCR products confirmed the intended PMMoV target region in all products (Table S8), further confirming the assay specificity.

For *Cryptosporidium*, RT-nRPA-CONAN yielded positive results in 11 of 23 samples, whereas RT-nRPA-DETECTR yielded seven positive results. In contrast, substantially higher numbers of positive calls were obtained using RT-LAMP and the two qPCR assays, with positive signals observed in 19 samples by RT-LAMP, 21 samples by qPCR-NJ, and all 23 samples by qPCR-CM (Table S3). However, nanopore sequencing demonstrated that the higher apparent positive rates of these assays did not necessarily coincide with amplification of *Cryptosporidium*-specific sequences. To evaluate whether each assay result reflected amplification of the intended target, the presence or absence of *Cryptosporidium*-specific contigs in the corresponding amplification products was used as the reference for calculating assay sensitivity and specificity. For RT-nRPA-CONAN, all 11 samples containing *Cryptosporidium*-specific contigs yielded positive LFA signals, whereas the others 12 samples, which tested negative, lacked *Cryptosporidium*-specific contigs. Consequently, RT-nRPA-CONAN showed a sensitivity of 100% [95% CI: 76–100%] and a specificity of 100% [95% CI: 76–100%] (Figure 4C, D). RT-nRPA-DETECTR likewise showed a specificity of 100% [95% CI: 76–100%] but yielded positive results in only seven of the 11 samples containing *Cryptosporidium*-specific contigs, leading to a sensitivity of 64% [95% CI: 35–85%]. In the four DETECTR false-negative samples, *Cryptosporidium*-specific contigs were recovered from the corresponding nRPA products, indicating that DETECTR did not generate an LFA signal distinguishable from the negative control. In these samples, *Cryptosporidium*-specific reads accounted for 0.5-1.5% of the total nanopore reads, suggesting that the target amplicons constituted only a minor fraction of the amplified products, consistent with the weak LFA signals observed.

In contrast, LAMP and both qPCR assays frequently yielded positive signals, even when no *Cryptosporidium*-specific contigs were recovered from the amplification products. Nanopore sequencing revealed that several of these false-positive products instead contained sequences derived from non-target environmental protists or diatoms. Specifically, LAMP products contained sequences matching e.g., *Dinophysis* sp. and *Anteholosticha* sp., qPCR-NJ products contained sequences matching e.g., *Salpingoeca* sp., and qPCR-CM products contained sequences matching e.g., *Nitzschia* sp. These findings indicate that nonspecific amplification of environmental eukaryotic nucleic acids contributed to false-positive detection by these assays. Accordingly, although their sensitivities were >88%, their specificities were lower, at 30% [95% CI: 11–60%] for RT-LAMP, 6.7% [95% CI: 1.2–30%] for qPCR-NJ, and 0% [95% CI: 0–26%] for qPCR-CM, which were significantly lower than those of RT-nRPA-CONAN and RT-nRPA-DETECTR (McNemar’s test, *p* < 0.05).

Overall, RT-nRPA-CONAN showed the highest sensitivity and specificity among the evaluated assays, indicating that combining isothermal amplification with CRISPR-mediated sequence recognition improves the specificity of *Cryptosporidium* detection in complex environmental water samples.

## 3. Discussion

For environmental water, where target concentrations can be low and only a small fraction of nucleic acid is subjected to molecular assays, improving analytical LOD is mandatory for practical biomarker monitoring in water. The present study demonstrates that optimizing the upstream amplification process can lower the analytical LOD of LFA-based CONAN. The major bottleneck was the inability of conventional RPA to generate sufficient amplicon concentrations from low-copy RNA inputs. Introducing a nested RPA step increased the amplification capacity and lowered the analytical LOD of RT-nRPA-CONAN to the few-copy-per-reaction range.

The nRPA-CONAN approach also showed an advantage in specificity when applied to environmental water samples. Although the qPCR and LAMP assays used here had previously been developed for *Cryptosporidium* detection, nanopore sequencing revealed that several apparent positive reactions did not contain *Cryptosporidium*-specific contigs but instead contained sequences derived from non-target environmental eukaryotes. False-positive *Cryptosporidium* detection using TaqMan-based qPCR assays has likewise been reported in environmental water samples^30,31^. For LAMP, false-positive reactions has not specifically tested for this *Cryptosporidium* assay but have been reported for other microbial targets, emphasizing the need for validation across different matrices and empirical optimization of reaction time^32,33^. More broadly, they highlight an important challenge in traditional molecular assays for environmental water monitoring; assays validated using defined panels of target and non-target organisms may not necessarily retain the same specificity when applied to natural waters containing much greater taxonomic and sequence diversity. Given that even nRPA products contained heterogeneous amplicon populations (Table S4), an additional layer of sequence recognition by CRISPR-Cas protein is essential for specific identification of *Cryptosporidium* 18S rRNA. It should be noted, however, that all PMMoV-positive reactions coincided with the presence of intended targets for both qPCR and nRPA-CONAN, suggesting that assay performance of qPCR in environmental waters also depends on the target and sample context.

There are important practical and analytical trade-offs among nRPA-CONAN, LAMP, and qPCR as summarized in Table 3. Notable advantages of nRPA-CONAN include its low instrumentation requirements and intuitive visual readout, together with a low analytical LOD and high specificity. A previous study demonstrated that both RPA and CONAN reactions can be performed using an electricity-free hand warmer as a heat source, highlighting the potential of this platform for use in resource-limited settings^16^. In contrast, assay design remains less straightforward than for qPCR because substantial empirical screening of RPA primers and optimization of crRNA design may be required. Analogous to CONAN-SWIFT for computational design of LAMP-CONAN assays^28^, RPA-specific primer-design tools could facilitate the development and broader adoption of RPA-coupled CRISPR diagnostics.

**Table 3.** Comparison of practical and analytical characteristics of nRPA-CONAN, LAMP, and qPCR for RNA-target detection. Reported sensitivity and specificity values were obtained from the environmental samples examined in this study, using the presence or absence of *Cryptosporidium*-specific contigs in the corresponding amplification products as the assay-specific reference.

| Feature | nRPA-CONAN | LAMP | qPCR |
|---|---|---|---|
| Number of primers | 3 or 4 | 4–6 | 2 |
| Probe / crRNA | At least 1 crRNA | None | At least 1 probe |
| Design constraints | PAM and crRNA sequence/length constraints | Multiple primer-site and spacing constraints | Primer/probe sequence and melting temperature constraints |
| Availability of primer/probe/crRNA design tools | Limited; Empirical screening required | Yes<sup>28</sup>; Empirical screening required | Yes<sup>41</sup>; highly established. |
| Reaction temperature | 37°C | 60–65°C | Thermal cycling |
| RNA detection limit | Few copies | Typically, few copies | Few copies |
| Detection format | Test strip; fluorescence | Fluorescence; turbidity; colorimetric; test strip | Fluorescence |
| Reaction configuration | Two-pot | One-pot | One-pot; Two-pot |
| Quantification | Primarily qualitative | Primarily qualitative | Quantitative |
| Specificity in this study | 100% | 30% | 0-6.7% |
| Sensitivity in this study | 100% | 92% | 88-100% |
| Special equipment required | Heater | Heater; Fluorescence or turbidity meter | Thermal cycler with fluorescence reader |

Instrument-light nucleic acid detection could expand microbial testing capacity by reducing the resources required per sample. This may be particularly relevant for *Cryptosporidium* monitoring, for which conventional molecular workflows often involve concentration, immunomagnetic separation (IMS), nucleic-acid extraction, and qPCR. In the present study, *Cryptosporidium* can be detected without IMS or a thermal cycler, reducing the equipment requirements and costs associated with detection. Potential resource savings could support more intensive sampling, including higher-frequency or event-triggered monitoring during rainfall or other contamination events, as increasingly advocated for drinking-water source protection^34^. Currently, Japanese drinking-water guidelines recommend monitoring of *Cryptosporidium* more than once every three months and daily collection and 14-day storage of 20-L finished-water samples for higher-risk facilities^35^. For example, one potential strategy to optimize testing capacity would be increasing sampling frequency and introducing CONAN as a screening tool, where only CONAN-positive samples can be prioritized for microscopy-based confirmation and enumeration, where required. Further simplification of sample processing (e.g., field-deployable extraction) will be essential for adapting the current thermal-cycler-free assay to decentralized water monitoring^36^ and WBE^37^. Such an approach could not only reduce the resources required per test but also enable more flexible monitoring frameworks in resource-limited settings.

This study has several limitations that should be addressed in future studies. First, the present study optimized only CONAN, whereas LAMP and qPCR followed published protocols; their observed performance should not be interpreted as the maximum achievable for those technologies. Second, the environmental validation comprised 23 river water and wastewater samples and should be extended across matrices, sites and extraction workflows. Finally, cross-study comparison of detection limits for CRISPR diagnostics still remains difficult because molar or concentration-based LODs are often reported on different volume bases^38^. For example, a previous RPA-Cas12a-based assay reported an LOD of 6.88 aM for *Vibrio* DNA detection, though the denominator was the reaction volume of RPA, 50 µL^10^, and thus the LOD was actually equivalent to 207 copies/reaction of RPA. Another recombinase-assisted amplificaition-Cas12a assay reported a sensitivity of 0.1 copy/µL for detecting norovirus GII.4 and GII.17^39^ without describing the volume basis used for the reported concentration. Future studies should report analytical LODs as target copies introduced into the initial amplification reaction and compare assays using identical target inputs, with template and reaction volumes explicitly stated^40^.

## 4. Conclusions

This study developed a highly sensitive and specific RT-nRPA-CONAN workflow for test-strip-based detection of *Cryptosporidium* 18S rRNA and PMMoV in environmental water samples. By introducing nested RPA to overcome insufficient amplification by conventional RPA, the CONAN assay achieved a few-copy analytical limit of detection while retaining a lateral-flow readout. Application to river water and wastewater extracts further showed that CRISPR-mediated sequence recognition can enhance specificity by distinguishing intended target amplicons from nonspecific environmental amplicons. The nRPA-CONAN workflow provides a basis for decentralized and instrument-light monitoring of nucleic acid biomarkers and highlights the potential of CRISPR diagnostics as a screening for health-related water microorganisms where rapid qualitative detection with high specificity is required.

## 5. Methods

### 5.1. Protein, DNA, and crRNA

Cas3 and 2A Cascade protein were synthesized and purified as described elsewhere.^42^ Alt-R LbCas12a was purchased from Integrated DNA Technologies (IDT).

All the primers, probes, and crRNAs were synthesized and purchased from FASMAC, IDT, or GenScript. Representative 75% consensus target sequences of different *Cryptosporidium* species were synthesized as gBlocks Gene Fragments (IDT). Total RNA of each organism was extracted from viable *C. parvum* purchased from Waterborne Inc. and pepper mild mottle virus (MAFF #104032, NARO GEnebank). RNA concentrations were measured by droplet digital PCR as described in the SI.

### 5.2. CRISPR diagnostics

#### 5.2.1. RT-nested RPA (RT-nRPA)

RT-nested RPA (RT-nRPA) was performed in a one-pot reaction with a final volume of 25 μL. Each lyophilized pellet was resuspended with 29.5 μL of rehydration buffer (TwistDx).

First, 2.95 μL of the rehydrated RPA mastermix was supplemented with 0.25 μL of outer primer mix (F1 and R1; each at 10 μM), 20 U of SuperScript IV reverse transcriptase (Thermo Fisher Scientific), 1 U of RNase H (NEB), and 1 μL of template to a total volume of 4.5 μL. The mixture was incubated at 37 °C for 10 min to allow reverse transcription. Note that the inclusion of RNase H has been reported to lower the analytical LOD by >10-fold, and the same effect was observed under our experimental conditions (data not shown) likely because RNase H selectively degrades the RNA strand of the RNA:cDNA hybrid, thereby making the resulting DNA strand accessible to recombinase^43,44^.

The first RPA reaction was then initiated by adding 0.5 μL of 140 mM magnesium acetate, followed by incubation at 37 °C for an additional 10 min.

Subsequently, 18 μL of the second RPA reaction mix, containing 11.8 μL of RPA mastermix, 4.2 μL of nuclease-free water, and 2 μL of inner primer mix (F2 and R2; each at 10 μM), was added to the reaction. The second RPA reaction was initiated by adding 2 μL of 140 mM magnesium acetate, followed by incubation at 37 °C for an additional 15 min. The outer primers were used at half the concentration of the inner primers; combined with the addition of 20 µL of second-round mixture to the 5-µL first-round reaction, this resulted in an eightfold higher final concentration of inner primers than outer primers during the second RPA reaction, favoring amplification by the inner primers. RT-nRPA products were immediately subjected to either CONAN or DETECTR assays or stored at −20°C until processing.

#### 5.2.2. CONAN

CONAN reactions were performed in a total volume of 10 μL with slight modifications to a previous report^16^. Prior to the assay, 0.4 mg/mL of 2A-Cascade was preincubated with 1 μM of crRNA for 10 min at 37 °C. The Cascade-crRNA complex was used immediately or stored at 4 °C for up to 8 h before use. Each reaction contained 100 nM Cascade-crRNA complex, 400 nM Cas3, 4.7 mM ATP, and CRISPR-Cas3 system working buffer (60 mM KCl, 10 mM MgCl_2_, 10 μM CoCl_2_, 5 mM HEPES-KOH pH 7.5), 1 μL of nRPA products, and 1 μL of 5 nM cleavage reporter (FQ probe:/56FAM/ATATAT/3BHQ1/ for fluorescence assay, FB probe:/56FAM/ATATAT/3Biotin/; for lateral-flow assay; FASMAC). For positive and negative controls, 10^3^ copies of 18S rRNA of *C. parvum* and nuclease-free water were included.

For fluorophore quencher (FQ)-labeled reporter assay, the reactions were incubated in a thermal cycler (QuantStudio 5, Thermo Fisher Scientific) for up to 20 min at 37 °C with fluorescence measured every 30 seconds (λ_ex_: 470 nm, λ_em_: 520 nm). Changes in the fluorescence values were analyzed by QuantStudioTM Design & Analysis Software v1.5.2 (Thermo Fisher Scientific). The initial fluorescence increase rate was calculated as the slope of the linear regression of fluorescence values measured between 0.5 and 2.5 min.

For the lateral flow assay, reactions were incubated for 10 min at 37°C, after which the entire reaction mixture was applied to the sample application pad of a lateral-flow strip (Milenia HybriDetect 1; TwistDx) and dipped into 80 μL of dipstick assay buffer. Results were visually assessed after approximately 2 min. A single band near the sample application pad was interpreted as a negative result, whereas a single band near the top of the strip or two bands were interpreted as a positive result (Figure 1). Images were converted to 8-bit format, and band intensities were quantified using ImageJ. The mean intensity of each band was normalized to the signal intensity of an adjacent background region. Band intensities more than 0.99 quantile of non-template control for each assay were regarded as positive.

#### 5.2.3. DETECTR

DETECTR reactions were performed according to a previous report^12^, in a total volume of 10 μL. Prior to the assay, 62.5 nM LbCas12a (IDT) was preincubated with 78.1 nM of crRNA in 1.25x NEBuffer r2.1 for 20 min at 37°C to allow formation of the crRNA-LbCas12a complex. The RNA-protein complex was used immediately or stored at 4 °C for up to 8 h before use.

For each reaction, 8 μL of the RNA-protein complex was mixed with 1 μL of RPA product and 1 μL of the same 5 nM cleavage reporter used for CONAN. For positive and negative controls, 10^3^ copies of 18S rRNA of *C. parvum* and nuclease-free water were included. Both the FQ reporter assay and LFA were performed in the same manner as described for CONAN.

### 5.3. Traditional molecular methods

#### 5.3.1. RT-LAMP

RT-LAMP reactions were performed in a total volume of 12.5 μL, which consisted of WarmStart LAMP 2X Master Mix, 50x Fluorescent dye (WarmStart LAMP Kit (DNA & RNA), NEB), 10x LAMP primer mix, according to a previous study^21^, and 1 μL of nucleic acid extracts. For positive and negative controls, 10^3^ copies of 18S rRNA of *C. parvum* and nuclease-free water were included. Reactions were incubated in a thermal cycler for up to 50 min at 60 °C with fluorescence measured every 30 seconds (λ_ex_: 470 nm, λ_em_: 520 nm). Changes in the fluorescence values were analyzed by QuantStudioTM Design & Analysis Software v1.5.2 (Thermo Fisher Scientific).

#### 5.3.2. RT and qPCR

A 4 μL of the extract was subjected to reverse transcription (RT) using the High-Capacity cDNA Reverse Transcription Kit (Thermo Fisher Scientific) according to the manufacturer’s protocol, yielding 8 μL of cDNA.

Real-time PCR was performed in a total reaction volume of 25 μL, which consisted of 2 μL of the template, 2x QuantiTect Probe PCR Master Mix (Qiagen), 10 pmol each of the forward and reverse primer, and 5 pmol of probe. The primers and probes used in this study were those described in previous studies^22,24,25^. Reactions were performed under the following thermal conditions: 50 °C for 2 min and initial denaturation at 95 °C for 15 min, followed by 45 cycles of denaturation at 95 °C for 15 sec and annealing and extension at 60 °C for 1 min. For non-template control (NTC), nuclease-free water instead of template was included. Amplification data were analyzed by QuantStudio^TM^ Design & Analysis Software v1.5.2 (Thermo Fisher Scientific).

### 5.4. Sampling sites, sample collection, concentration, and extraction

A total of 17 river water samples were collected at sites within the Sagamigawa, Tamagawa, and Arakawa River basins in Japan between 2023 and 2026 as illustrated in Figure S5. These sampling sites were selected because they are influenced by municipal and/or livestock wastewater, and *Cryptosporidium* oocysts have been sporadically detected by immunofluorescence microscopy in the past prefectural surveillance. In addition, three wastewater samples each were collected from municipal wastewater treatment plants in Sapporo, Japan and an airport in Japan. All samples were transported to the lab under cool conditions and processed within 1 day of collection.

Concentration and nucleic acid extraction were performed according to previously reported methods.^31,45^ Briefly, 1-2 L of river water or 300 mL of wastewater was supplemented with MgCl_2_ to a final concentration of 25 mM and filtered through a 90-mm-diameter mixed cellulose ester membrane filter with a pore size of 0.8 μm. The membrane filters were cut into four equal pieces and stored at −20°C. Nucleic acids were directly extracted from one quarter of each filter by bead-beating, followed by purification and elution in 50 μL of DNase/RNase-free water. This workflow was reported to simultaneously concentrate and extract *C. parvum* rRNA and a virus target, murine norovirus, with >83% recovery^31^.

### 5.5. Bioinformatics and statistical analysis

The 18S rRNA sequences of multiple *Cryptosporidium* species were retrieved from GenBank and aligned. A 90% consensus sequence was generated for each of *C. parvum, C. hominis, C. meleagridis, C. felis, C. canis, C. muris, C. andersoni, C. avium, C. suis, C. bovis*. For PMMoV, a total of 78 near-full-length genome sequences were retrieved from GenBank and aligned. The alignment was performed with MAFFT.

Nanopore reads were trimmed using the corresponding primer sequences and de novo assembled, and contigs aligned to the intended target region with 100% sequence identity across the amplified target region were classified as *Cryptosporidium*-specific or PMMoV-specific using Geneious Prime 2025.3.1.

The input copy number corresponding to a 50% probability of detection (LOD_50_) was estimated with a single-hit Poisson model.

## Data Availability

All data produced in the present work will be deposited in Zenodo upon acceptance.

## Acknowledgement

This study was supported by AMED under Grant Numbers, JP223fa627001, JP223fa627001 (UTOPIA Next-Generation Diagnostic Tool Discovery Program), JP223fa727002, 26ck0106096h and 26fk0108713h0003, the River Fund of The River Foundation, Japan under Grant Number 2023-5311-005, Kurita Water and Environmental Foundation, the Japan Society for the Promotion of Science (JSPS) through Grant-in-Aid for Scientific Research (A) (grant numbers JP23H00367 and JP24H00326), Grant-in-Aid for Scientific Research (B) (grant numbers JP24K02010 and JP26K01072), the Environment Research and Technology Development Fund (grant numbers JPMEERF20255RB1 and JPMEERF20265001) of the Environmental Restoration and Conservation Agency provided by Ministry of the Environment of Japan. During the preparation of this work, the authors used GPT-5.6 Sol (OpenAI) and Claude Science (Anthropic, Claude Opus 5.5 model) for proofreading the manuscript and for assisting with R code development. After using these tools, the authors reviewed and edited the content as needed and take full responsibility for the content of the publication.

## Notes

### Competing Interest Statement

T.M. and K.Y. serve as scientific advisors to C4U Corporation, which holds patents related to the CONAN technology.

## References

1. Yang, Z. Low-cost and rapid sensors for wastewater surveillance at low-resource settings. Nat. Water 1, 405–407 (2023).

2. Notomi, T. et al. Loop-mediated isothermal amplification of DNA. Nucleic Acids Res. 28, e63–e63 (2000).

3. Piepenburg, O., Williams, C. H., Stemple, D. L. & Armes, N. A. DNA Detection Using Recombination Proteins. PLOS Biol. 4, e204 (2006).

4. Cao, H. et al. Paper Device Combining CRISPR/Cas12a and Reverse-Transcription Loop-Mediated Isothermal Amplification for SARS-CoV-2 Detection in Wastewater. Environ. Sci. Technol. 56, 13245–13253 (2022).

5. Lobato, I. M. & O’Sullivan, C. K. Recombinase polymerase amplification: Basics, applications and recent advances. TrAC Trends Anal. Chem. 98, 19–35 (2018).

6. Shirato, K. et al. Development of fluorescent reverse transcription loop-mediated isothermal amplification (RT-LAMP) using quenching probes for the detection of the Middle East respiratory syndrome coronavirus. J. Virol. Methods 258, 41–48 (2018).

7. Huang, Y.-H. & Jiang, S. Quantification of Viruses in Wastewater on a Centrifugal Microfluidic Disc. Environ. Sci. Technol. 59, 3088–3097 (2025).

8. Kaminski, M. M., Abudayyeh, O. O., Gootenberg, J. S., Zhang, F. & Collins, J. J. CRISPR-based diagnostics. Nat. Biomed. Eng. 5, 643–656 (2021).

9. Cheng, Z.-H. et al. WATER NEWS: a field approach for sustainable detection of pathogens in wastewater. Natl. Sci. Rev. 12, nwaf275 (2025).

10. Kim, N. et al. A field-deployable CRISPR-based biosensing platform for monitoring marine ecosystems. Nat. Sustain. 9, 51–64 (2026).

11. Sun, Z. et al. A contamination-free lateral flow platform for point-of-care virus detection in wastewater. Environ. Chem. Lett. 10.1007/s10311-026-01924-0 (2026) doi:10.1007/s10311-026-01924-0.

12. Chen, J. S. et al. CRISPR-Cas12a target binding unleashes indiscriminate single-stranded DNase activity. Science 360, 436–439 (2018).

13. Yoshimi, K. et al. Dynamic mechanisms of CRISPR interference by Escherichia coli CRISPR-Cas3. Nat. Commun. 13, 4917 (2022).

14. Larremore, D. B. et al. Test sensitivity is secondary to frequency and turnaround time for COVID-19 screening. Sci. Adv. 7, eabd5393 (2021).

15. Wang, S. & Hasan, R. CRISPR Biosensing for Environmental Monitoring: Workflow Design and Performance Benchmarking. Environ. Sci. Technol. 10.1021/acs.est.6c02006 (2026) doi:10.1021/acs.est.6c02006.

16. Hirano, R. et al. Sustainable and portable CRISPR-based diagnostics for high-sensitivity Mpox detection. Npj Biosensing 2, 43 (2025).

17. Hamilton, K. A. et al. Cryptosporidium and Giardia in Wastewater and Surface Water Environments. J. Environ. Qual. 47, 1006–1023 (2018).

18. Kitajima, M., Sassi, H. P. & Torrey, J. R. Pepper mild mottle virus as a water quality indicator. Npj Clean Water 1, (2018).

19. Yoshimi, K., et al. CRISPR-Cas3-based diagnostics for SARS-CoV-2 and influenza virus. iScience 25, (2022).

20. Crannell, Z. A. et al. Nucleic Acid Test to Diagnose Cryptosporidiosis: Lab Assessment in Animal and Patient Specimens. Anal. Chem. 86, 2565–2571 (2014).

21. Inomata, A. et al. Development and evaluation of a reverse transcription-loop-mediated isothermal amplification assay for rapid and high-sensitive detection of Cryptosporidium in water samples. Water Sci. Technol. 60, 2167–2172 (2009).

22. Jothikumar, N., da Silva, A. J., Moura, I., Qvarnstrom, Y. & Hill, V. R. Detection and differentiation of Cryptosporidium hominis and Cryptosporidium parvum by dual TaqMan assays. Journal of Medical Microbiology vol. 57 1099–1105 (2008).

23. Liu, Y. et al. Rapid detection of Cryptosporidium spp. in diarrheic cattle feces by isothermal recombinase polymerase amplification assays. Heliyon 9, (2023).

24. Mary, C. et al. Multicentric Evaluation of a New Real-Time PCR Assay for Quantification of Cryptosporidium spp. and Identification of Cryptosporidium parvum and Cryptosporidium hominis. J. Clin. Microbiol. 51, 2556–2563 (2013).

25. Wang, D. et al. Novel strategy to quantify the viability of oocysts of *Cryptosporidium parvum* and *C. hominis*, a risk factor of the waterborne protozoan pathogens of public health concern. Water Res. 258, 121788 (2024).

26. Haramoto, E. et al. Occurrence of pepper mild mottle virus in drinking water sources in Japan. Appl. Environ. Microbiol. 79, 7413–7418 (2013).

27. Zhang, T. et al. RNA viral community in human feces: Prevalence of plant pathogenic viruses. PLoS Biol. 4, 0108–0118 (2006).

28. Nakamura, J. et al. A rapidly deployable CRISPR–Cas3 diagnostic platform for emerging RNA viruses. 2026.08.25.746999 Preprint at 10.64898/2026.08.25.746999 (2026).

29. Hasan, R., Cragg, M. A. & Wang, S. A Portable RPA-CRISPR/Cas12a-Based Biosensing Platform for On-Site Detection of the Microcystin Synthetase E Gene in Lake Water. ACS EST Eng. 5, 3479–3488 (2025).

30. Staggs, S. E. et al. The Applicability of TaqMan-Based Quantitative Real-Time PCR Assays for Detecting and Enumerating Cryptosporidium spp. Oocysts in the Environment. PLOS ONE 8, e66562 (2013).

31. Torii, S. et al. Simultaneous detection of protozoa, bacteria, and viruses from environmental water through membrane-adsorption followed by direct nucleic acid extraction. Water Res. 298, 125792 (2026).

32. Kang, S., Zimmermann, M., Reinhart, M. & Julian, T. R. Rapid and quantitative loop-mediated isothermal amplification (LAMP) assays for discriminatory detection of Vibrio cholerae. Environ. Sci. Water Res. Technol. 12, 1222–1234 (2026).

33. Zimmermann, M., Schuppler, M., Julian, T. R. & Kang, S. Development of loop-mediated isothermal amplification (LAMP) assays for the detection of diarrheagenic E. coli in wastewater. Appl. Environ. Microbiol. 91, e00880–25 (2025).

34. Hachad, M. et al. β-D-glucuronidase activity triggered monitoring of fecal contamination using microbial and chemical source tracking markers at drinking water intakes. Water Res. 254, 121374 (2024).

35. Ministry of Health, Labour and Welfare, Japan. Guidelines for Cryptosporidium and Giardia Control in Water Supply. https://www.mlit.go.jp/common/830005030.pdf (2019).

36. Park, M. J. et al. Self-powered floating capsule for decentralized water detection and disinfection. Nat. Water 4, 895–904 (2026).

37. Chang, C.-L. et al. Wastewater intelligence predicts the emergence of clinically-relevant and drug-resistant Candidozyma auris at healthcare facilities. Nat. Commun. 17, 5415 (2026).

38. Santiago-Frangos, A. et al. CRISPR-Cas, Argonaute proteins and the emerging landscape of amplification-free diagnostics. Methods 205, 1–10 (2022).

39. Qian, W. et al. Ultrasensitive and visual detection of human norovirus genotype GII.4 or GII.17 using CRISPR-Cas12a assay. Virol. J. 19, 150 (2022).

40. Soh, J. H. et al. CRISPR-based systems for sensitive and rapid on-site COVID-19 diagnostics. Trends Biotechnol. 40, 1346–1360 (2022).

41. Untergasser, A. et al. Primer3—new capabilities and interfaces. Nucleic Acids Res. 40, e115 (2012).

42. Asano, K. et al. CRISPR Diagnostics for Quantification and Rapid Diagnosis of Myotonic Dystrophy Type 1 Repeat Expansion Disorders. ACS Synth. Biol. 13, 3926–3935 (2024).

43. Feng, W. et al. Integrating Reverse Transcription Recombinase Polymerase Amplification with CRISPR Technology for the One-Tube Assay of RNA. Anal. Chem. 93, 12808–12816 (2021).

44. Qian, J. et al. An enhanced isothermal amplification assay for viral detection. Nat. Commun. 11, 5920 (2020).

45. Ando, H. et al. Wastewater-based prediction of COVID-19 cases using a highly sensitive SARS-CoV-2 RNA detection method combined with mathematical modeling. Environ. Int. 107743 (2023).

