## Supplementary material for "CRISPR-Cas3-Based Detection of *Cryptosporidium* and Pepper Mild Mottle Virus in Environmental Waters Using a Test-Strip Readout": SI

### S1: *Cryptosporidium* and the Rationale for selecting 18S rRNA as a target.

*Cryptosporidium* spp. are protozoan enteric parasites that cause cryptosporidiosis and can be transmitted through contaminated drinking or recreational water[1]. Among the recognized species, *C. parvum* and *C. hominis* are the predominant species causing human cryptosporidiosis, while several other species, such as *C. meleagridis*, *C. felis* or *C. suis* can also infect humans[2]. Historically, *Cryptosporidium* in water has been detected primarily by microscopic examination, including immunofluorescent antibody staining. For water samples, conventional methods generally rely on concentration and immunomagnetic separation followed by immunofluorescence microscopy, as exemplified by US EPA Method 1623.

Molecular detection has increasingly been adopted as an alternative approach in both clinical and water-quality applications. For example, nucleic acid detection is accepted as laboratory evidence for confirmed cryptosporidiosis in clinical surveillance[3] and molecular methods are also used for concentrated water samples in some countries, including Japan[4]. These approaches can reduce reliance on microscopy-based immunofluorescent antibody test (IFAT), which is labor-intensive, costly and require special equipment and trained personnel.

Existing PCR methods target several genomic regions, including *Cryptosporidium* oocyst wall protein (COWP) gene and 18S rRNA gene[5] using either DNA or expressed RNA as the analytical target. Among these targets, 18S rRNA is particularly attractive for sensitive detection and was found to achieve sensitivity comparable to immunofluorescence microscopy[6] since a single viable oocyst contains >5,000 copies of 18S rRNA molecules, substantially more than other targets, with approximately 8-20 copies of 18S rDNA and a single copy of chromosomal gene such as COWP per oocyst. Given the low abundance of *Cryptosporidium* oocysts typically present in environmental water samples and the limited fraction of extracted nucleic acid that can be introduced into each molecular assay, 18S rRNA was therefore selected as the analytical target in this study.

### S2: Droplet digital PCR to enumerate RNA extracted from *C. parvum* and PMMoV

To quantify the RNA stocks used in this study, extracted RNA was diluted 1:100 and reverse-transcribed using the SuperScript™ IV First-Strand Synthesis System, with the same reverse transcriptase used for RT-(n)RPA. Droplet digital PCR (ddPCR) was performed in a total reaction volume of 20 μL, containing 2x ddPCR EvaGreen Supermix, forward and reverse primers at final concentrations of 250 nM, and 1 μL of cDNA template. *C. parvum* was quantified using forward primer 5′-GCGCAAATTACCCAATCCTAA-3′ (positions 413–433) and reverse primer 5′-GGGGTTTATACTTAACTCATTCC-3′ (positions 510–488), targeting a region within the C18S-F2/C18S-R2 amplicon, whereas PMMoV was quantified using the forward primer 5’-CTTGCATTGCAACCGACAATT- 3’, and reverse primer 5’- TGCTGCAACGACTCGATTTC - 3’. Droplets were generated using the QX200 Droplet Generator (Bio-Rad) and amplified using the

C1000 Touch Thermal Cycler (Bio-Rad). After amplification, droplets were analyzed using the QX200 droplet reader (Bio-Rad) and the Quantasoft Analysis Pro Software.

#### S3 Intercalator dye-based qPCR

Intercalator dye-based qPCR using SYBR Green was used exclusively to quantify *C. parvum* target concentrations in RPA products. RPA products were diluted 1:$10^2$ to 1:$10^6$ in nuclease-free water and used as templates. Reactions were performed in a total volume of 10 μL, containing 2x SsoAdvanced Universal SYBR Green Supermix (Bio-rad), the same primers as those described in Section S2, and 1 μL of template. A standard curve was generated using serial dilutions of the *C. parvum* gBlocks ($10^8$, $10^6$, $10^4$, $10^2$, $10^1$, gc/μL). For NTC, nuclease-free water instead of template was included. Specific amplification products were distinguished from primer dimers by analyzing the melting temperature of the positive control.

#### S4: Nanopore sequencing of amplified products

The nRPA, LAMP, and qPCR products were purified and submitted for nanopore sequencing using the Premium PCR sequencing service from Plasmidsaurus (USA). This service employs the ligation sequencing Kit v14 (Oxford Nanopore Technologies) and R10.4.1 flow cells (Oxford Nanopore Technologies), enabling an assembly accuracy of Q50 at a sequencing depth of 10–20× according to the service provider. A total of approximately 5,000 reads were obtained for each sample. Primer-derived sequences were trimmed before assembly: the inner forward and reverse primer sequences were removed from nRPA reads, FIP and BIP sequences were removed from LAMP reads, and the corresponding forward and reverse primer sequences were removed from qPCR reads. The trimmed reads were then de novo assembled, and the resulting contigs were aligned to the 18S rRNA gene of *C. parvum* strain C1 (GenBank accession no. AF108864) and PMMoV S strain (GenBank accession no. NC_003630) for screening the potential contigs matching the target sequences using Geneious Prime 2025.1.2. The resultant contigs were blasted and assessed to determine whether they are *Cryptosporidium*-specific or PMMoV-specific.

#### S5: Assessment of species coverage

To experimentally assess the species coverage of the designed CONAN assay, $10^4$ copies of synthetic dsDNA representing the consensus target sequences of *C. parvum*/*C. hominis*/*C. meleagridis, C. felis, C. canis, C. muris*/*C. andersoni*/*C. avium, C. suis,* and *C. bovis* were subjected to nRPA-CONAN followed by fluorescence and LFA readouts. Sequence alignment of the crRNA target regions among the tested *Cryptosporidium* species is shown in Figure S4A.

The fluorescence assay showed the strongest responses for *C. parvum*, *C. hominis*, *C. meleagridis*,

*C. felis*, and *C. suis* (Figure S4C, D). Reactivity toward *C. canis* and *C. bovis* was substantially lower, with 29-fold and 27-fold lower initial fluorescence increase rates, respectively, than those observed for the strongly reactive species. Only weak fluorescence responses were observed for *C. muris*, *C. andersoni*, and *C. avium*, with an initial fluorescence increase rate 69-fold lower than that of *C. parvum*/*C. hominis*/*C. meleagridis*. Consistent with the fluorescence results, LFA produced strong test bands for *C. parvum*, *C. hominis*, *C. meleagridis*, *C. felis*, and *C. suis*, whereas little or no detectable band formation was observed for *C. muris*, *C. andersoni*, and *C. avium*.

The species-dependent responses were consistent with the location and number of mismatches within the crRNA-target region. Notably, *C. felis*, which contained two mismatches toward the PAM-distal end of the target region, retained strong CONAN activity, indicating tolerance to a small number of distal mismatches. In contrast, *C. canis* showed a 29-fold reduction in initial fluorescence increase rate, and one of its mismatches was located 21 nt downstream of the PAM. This position lies outside the 20-nt target-recognition region typically used by Cas12a-based DETECTR but remains within the 32-nt recognition region of CONAN. Thus, the longer CONAN crRNA can utilize sequence variation located farther from the PAM than DETECTR. Increasing sequence divergence was associated with progressively lower collateral activity, as illustrated by the further reduction observed for *C. muris*/*C. andersoni*/*C. avium*.

Overall, these results suggest that increasing sequence divergence even at PAM-distal sequence reduces collateral cleavage and highlights the contribution of its extended crRNA-target recognition region to discrimination among closely related sequences. The assay showed strong reactivity toward the primary target species *C. parvum* and *C. hominis*, as well as *C. meleagridis, C. felis,* and *C. suis*, while showing substantially reduced reactivity toward the other tested *Cryptosporidium* species.

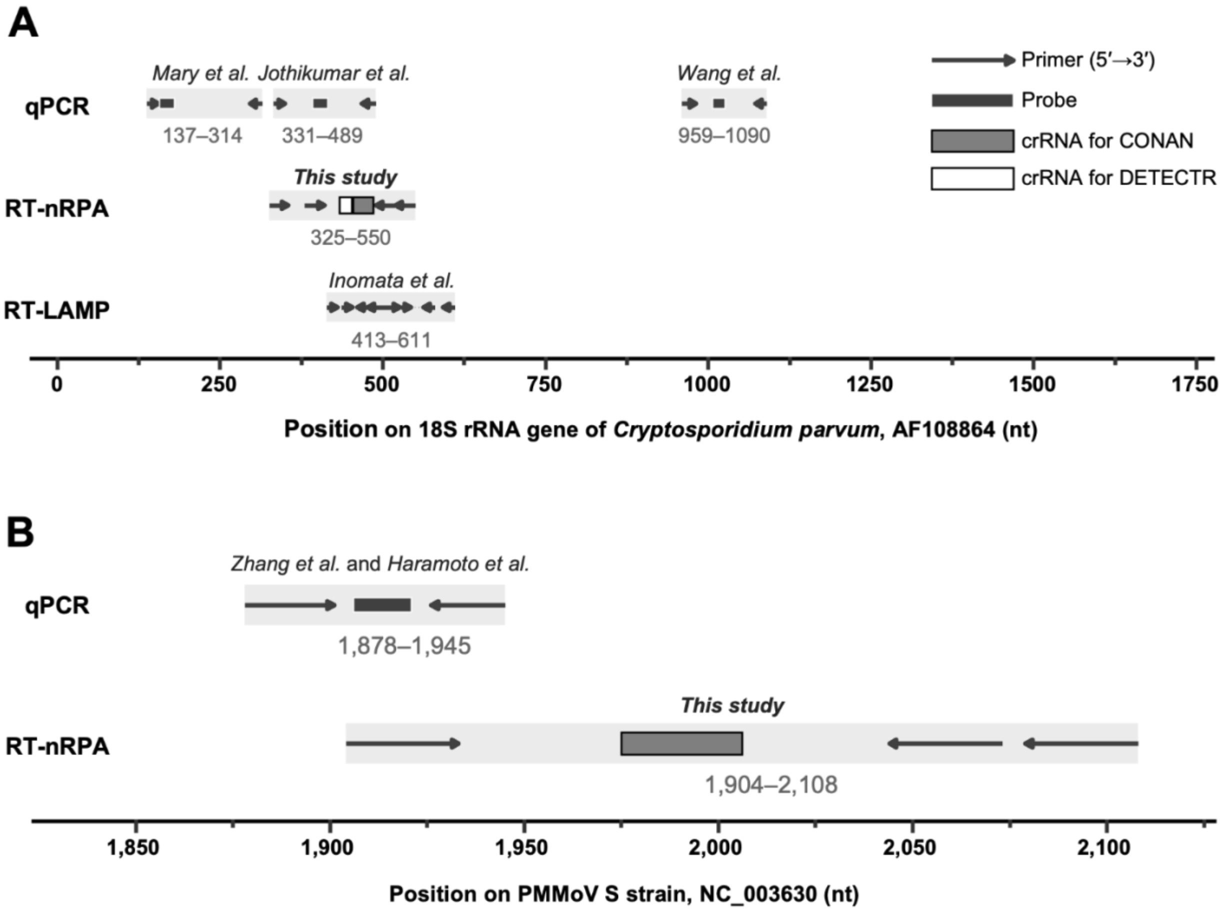


Figure S1 Locations of primers, probes, and crRNAs used for *Cryptosporidium* 18S rRNA (A) and PMMoV (B) assays. Arrows indicate primer-binding positions and their orientations. Numbers indicate nucleotide positions relative to the 18S rRNA gene of *Cryptosporidium parvum* strain C1 (GenBank accession no. AF108864) in (A) and the PMMoV S strain genome (GenBank accession no. NC_003630) in (B). Source studies are indicated above the shaded regions where applicable.

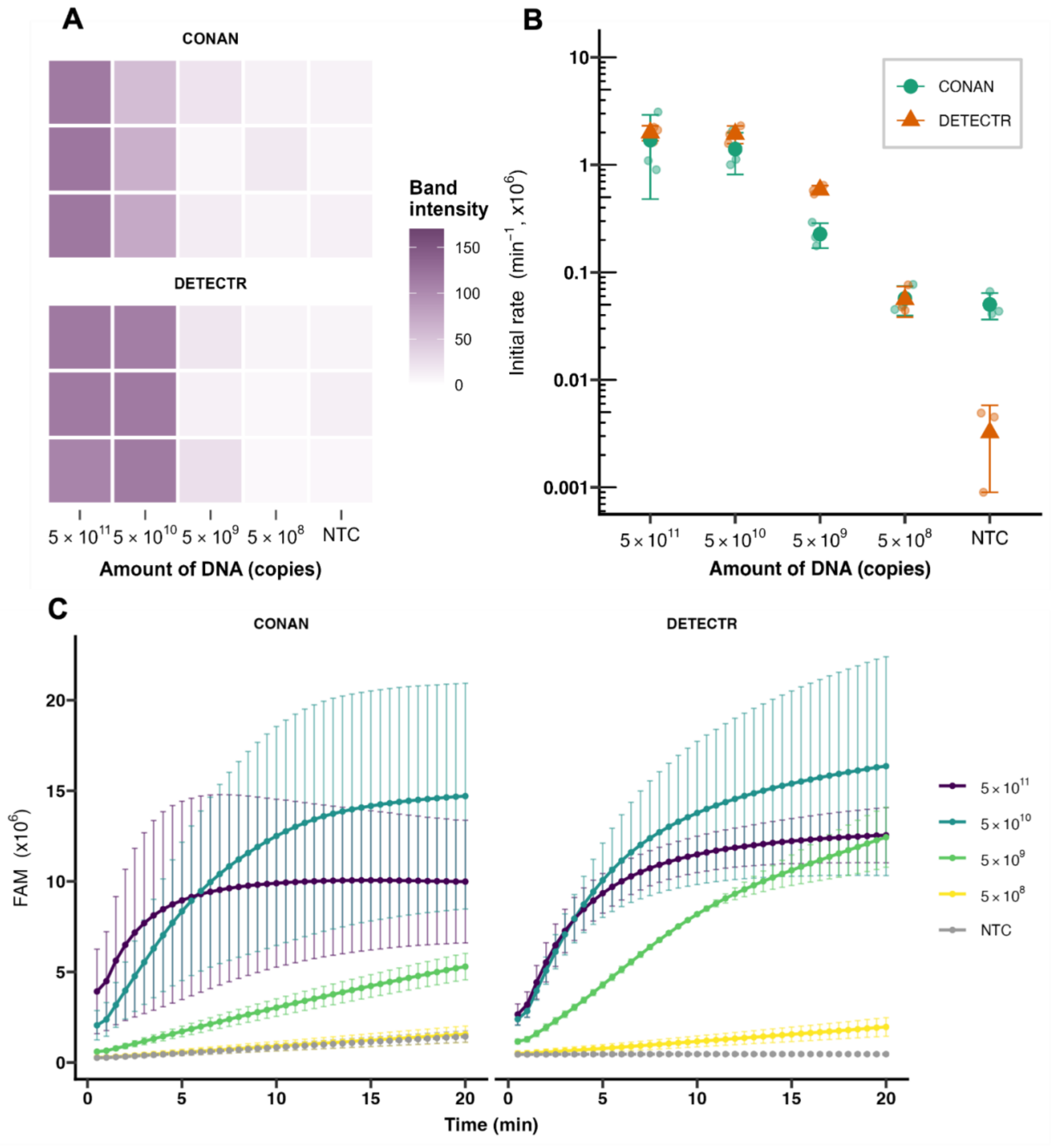


Figure S2 Comparison of CONAN and DETECTR responses across different DNA input levels. RT-nRPA products of *C. parvum* RNA were used as input. (A) Heatmaps of lateral-flow assay (LFA) test-band intensities for CONAN and DETECTR at DNA inputs, ranging from $5{\times}10^{11}$ to $5{\times}10^{8}$ copies and for the no-template control (NTC). Each row represents an independent replicate, and higher color intensity indicates a stronger test-band signal. (B) Initial fluorescence increase rates obtained with CONAN and DETECTR at the same DNA input levels. Small circle symbols indicate individual replicates, and large triangle symbols indicate the mean ± standard deviation. (C) Time-course fluorescence signals measured for CONAN and DETECTR over 20 min at each DNA input level. Data are shown as mean ± SD from independent replicates. NTC, no-template control.

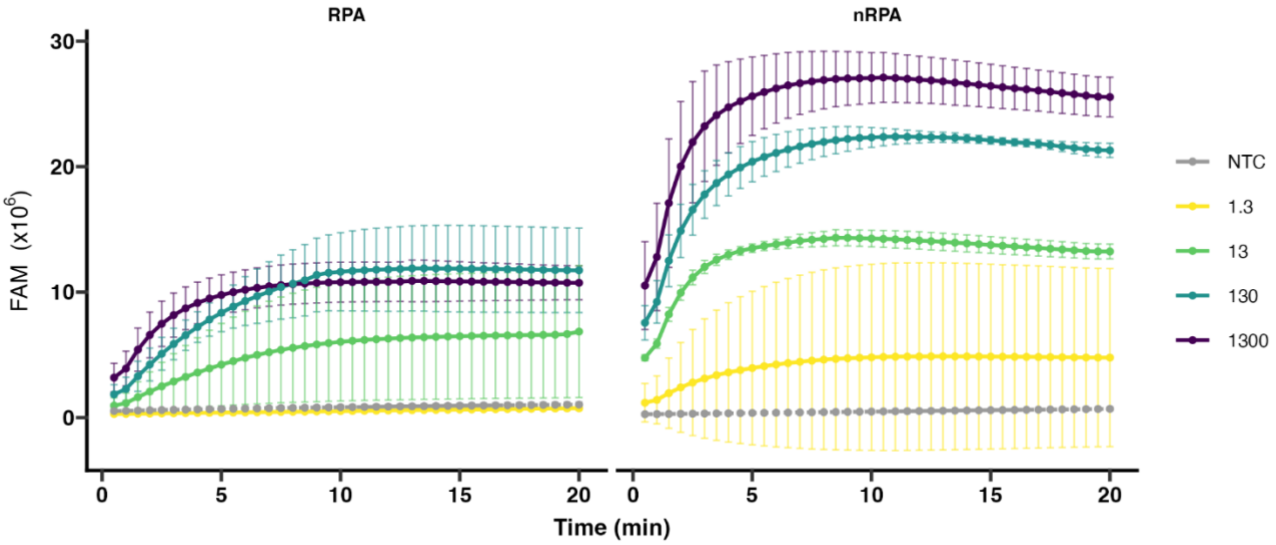


Figure S3 Time-course fluorescence responses of CONAN following conventional RPA or nested RPA (nRPA) at different initial RNA inputs. *C. parvum* 18S rRNA at different input levels was subjected to RT-RPA or RT-nRPA followed by CONAN, and fluorescence signals were monitored for 20 min. Data are shown as mean ± SD. NTC, no-template control.

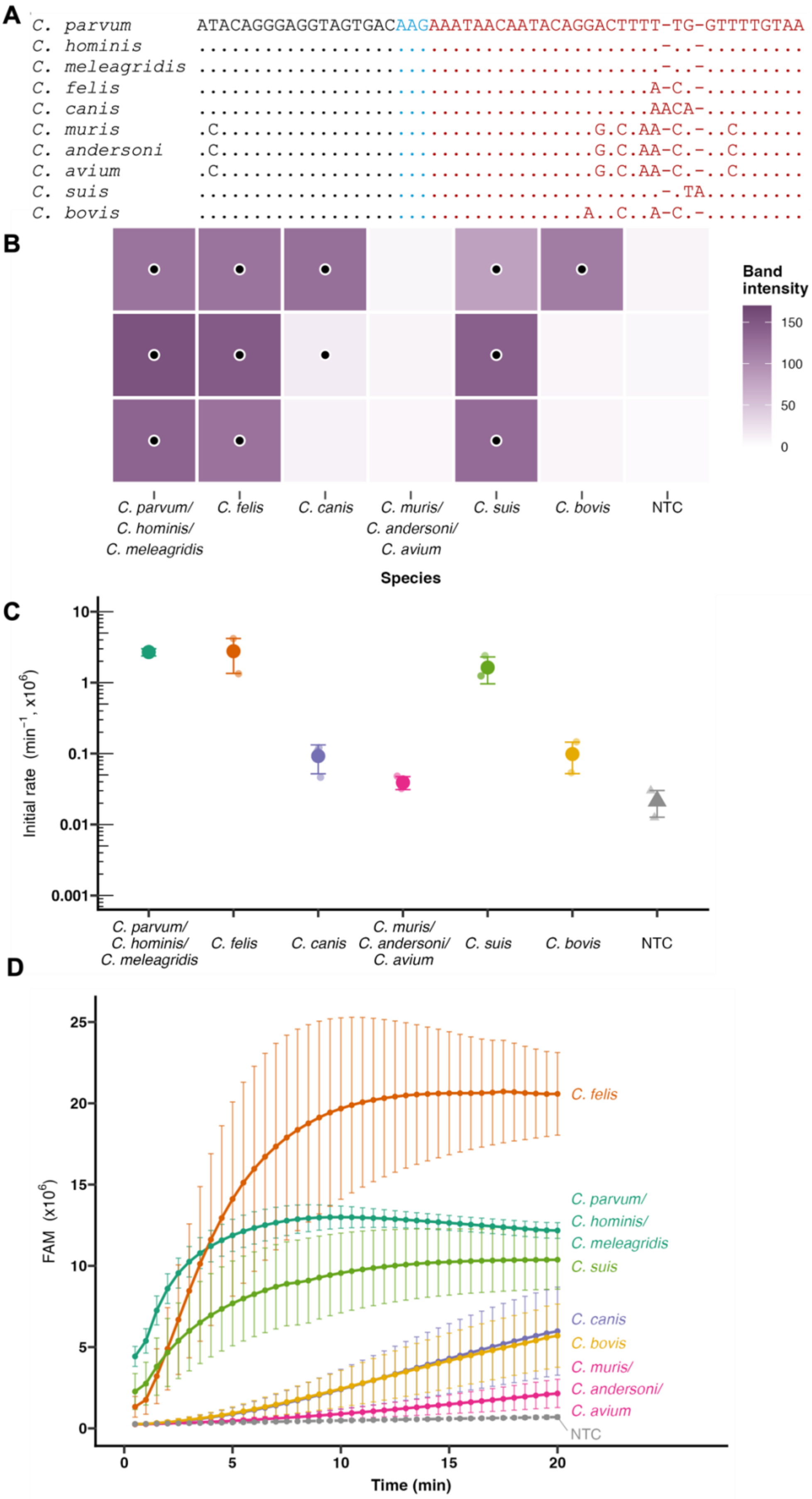
A
C. parvum ATACAGGGAGGTAGTGACAAGAAATAACAATACAGGACTTTT-TG-GTTTTGTAA
C. hominis
C. meleagridis
C. felis
C. canis
C. muris
C. andersoni
C. avium
C. suis
C. bovis
B
Band intensity
150
100
50
0
C. parvum/ C. hominis/ C. meleagridis
C. felis
C. canis
C. muris/ C. andersoni/ C. avium
C. suis
C. bovis
NTC
Species
C
Initial rate (min$^{-1}$, x10$^{6}$)
10
1
0.1
0.01
0.001
D
FAM (x10$^{6}$)
25
20
15
10
5
0
Time (min)
C. felis
C. parvum/
C. hominis/
C. meleagridis
C. suis
C. canis
C. bovis
C. muris/
C. andersoni/
C. avium
NTC

Figure S4 Species-dependent reactivity of the CONAN assay toward different *Cryptosporidium* species. (A) Sequence alignment of the crRNA target region among the tested *Cryptosporidium* species. The PAM and crRNA regions are shown in blue and red letters, respectively. Dots indicate nucleotides identical to the reference sequence, hyphens indicate gaps, and letters indicate nucleotide substitutions. (B) Heatmap of LFA test-band intensities obtained after nRPA-CONAN using $10^4$ copies of dsDNA representing the target sequences of different *Cryptosporidium* species. Black dots indicate samples classified as positive, with band intensities exceeding the upper 99% threshold of the non-template controls, calculated as the mean + 2.33 SD. NTC data were taken from the results of nRPA in Figure 3B for reference. (C) Initial fluorescence increase rates of the CONAN reaction for the tested *Cryptosporidium* species. Small symbols represent individual replicates, and large symbols represent the mean ± SD. (D) Time-course fluorescence signals of CONAN reactions for the tested *Cryptosporidium* species. Data are shown as mean ± SD. NTC, no-template control.

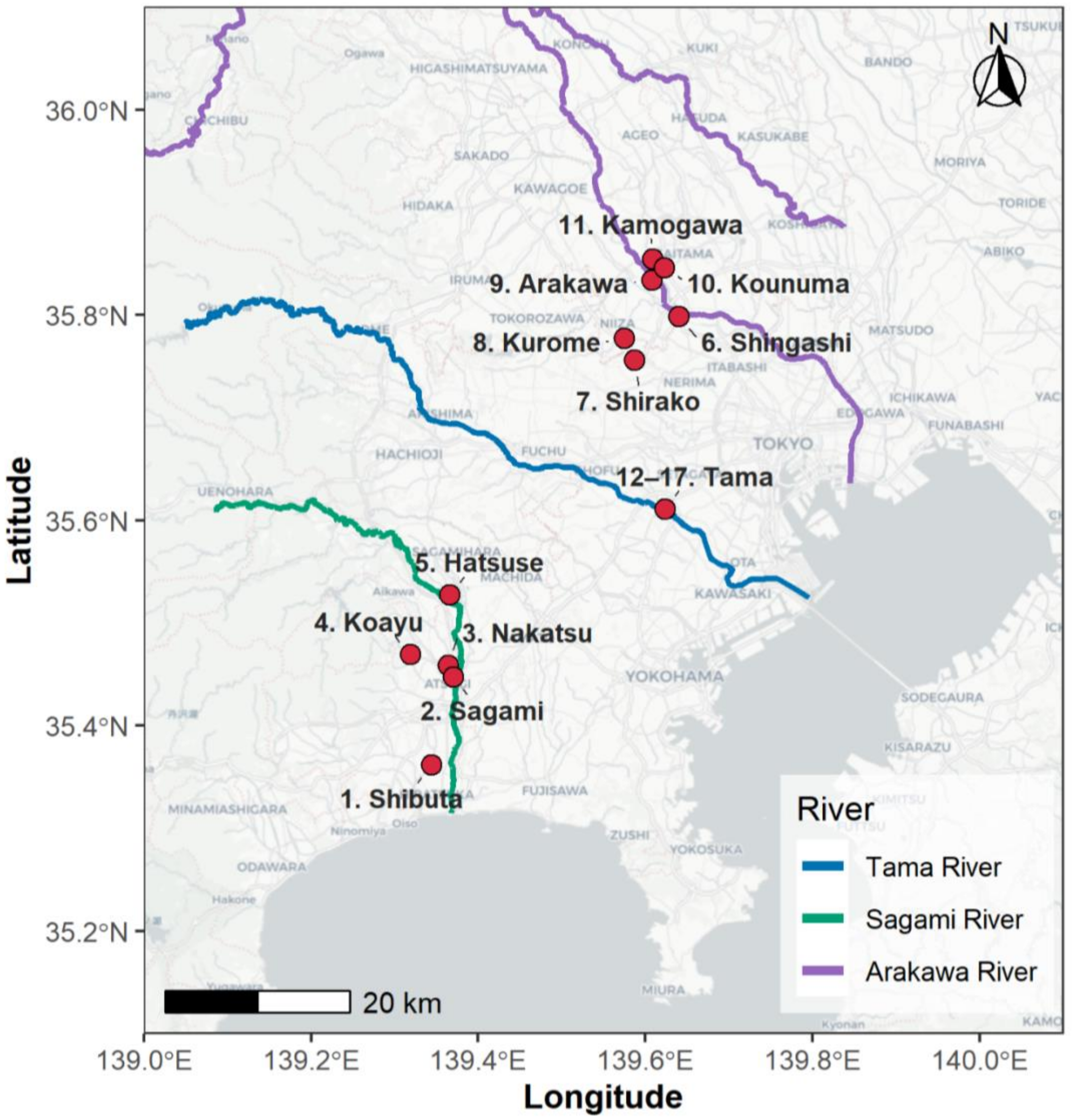


Figure S5 Sampling locations for river water samples collected from the Sagami, Tama, and Arakawa Rivers in Japan. Red circles indicate the sampling sites, and colored lines indicate the three river systems. The basemap is from © OpenStreetMap contributors, © CARTO. River network data are from © OpenStreetMap contributors, available under the Open Database License. Site names correspond to those listed in Table 2.

### References


1. Hamilton, K. A. *et al.* Cryptosporidium and Giardia in Wastewater and Surface Water Environments. *J. Environ. Qual.* **47**, 1006–1023 (2018).
2. Xiao, L. & Feng, Y. Molecular epidemiologic tools for waterborne pathogens *Cryptosporidium* spp. and *Giardia duodenalis*. *Food Waterborne Parasitol.* **8–9**, 14–32 (2017).
3. CDC. Clinical Testing and Diagnosis for Crypto. https://www.cdc.gov/cryptosporidium/hcp/diagnosis-testing/index.html (2025).
4. Ministry of Land, Infrastructure, Transport and Tourism. Testing Methods for Indicator Bacteria, Cryptosporidium, and Related Organisms in Water Supply No. 0330006. https://www.mlit.go.jp/common/830005142.pdf (2023).
5. USEPA. Selected Analytical Methods for Environmental Remediation and Recovery (SAM). https://www.epa.gov/esam/sam-pathogen-methods (2025).
6. Torii, S. *et al.* Simultaneous detection of protozoa, bacteria, and viruses from environmental water through membrane-adsorption followed by direct nucleic acid extraction. *Water Res.* **298**, 125792 (2026).
